# Validity and reproducibility of a culturally tailored dietary screening tool for hypertension risk in Nigerian healthcare

**DOI:** 10.1101/2024.01.24.24301732

**Authors:** Nimisoere P. Batubo, Carolyn I. Auma, J. Bernadette Moore, Michael A. Zulyniak

**Author notes:** Corresponding author (MAZ). **Author Contributions:**NPB and MAZ collaborated on the research methodology design. NPB led the trial, and NMN and CAA provided technical support. NPB led the analysis and prepared the first draft. MAZ and JBM provided analytical expertise. MAZ, JBM, and CIA provided critical feedback. NPB revised the manuscript. All authors approved the final manuscript.

## Abstract

Food frequency questionnaires (FFQ) are commonly used tools in dietary assessment but require validation. This study aimed to assess the relative validity and reproducibility of a culturally tailored FFQ for estimating food intake among Nigerian adults in clinical settings. The FFQ was administered to 58 patients at the Rivers State University Teaching Hospital, Nigeria, on two occasions, two weeks apart. Three repeat non-consecutive 24-hour dietary recalls (24DR) were also conducted as a reference method to evaluate the validity of the FFQ. Spearman’s rank correlations, Wilcoxon signed-rank tests, cross-classification agreement, intraclass correlation coefficients (ICC), and Bland-Altman analysis were performed in R to evaluate the relative validity and reproducibility. The trial was registered with *ClinicalTrials.gov:* NCT05973760. The correlation coefficient (*r*_s_) between the FFQ and 24DR ranged from 0.20 for ‘fats and oils’ to 0.78 for vegetables, with an average *r*_s_ of 0.60 (*p*<0.05). The Wilcoxon signed-rank tests indicated no significant differences in the 19 food groups queried (p>0.05), except for fats and oils (p<0.05). The exact agreement between FFQ and 24DR for classifying individuals into quartiles ranged from 17% for salt to 88% for processed meats and alcoholic drinks, with 90% of individuals classified into the same or neighbouring quartile. Additionally, the Bland-Altman analysis demonstrated acceptable agreement, with > 96% of observations within the acceptable limits of agreement for all food groups. For reproducibility, the ICC ranged from 0.31 for stew to 0.98 for fruit, with an average ICC of 0.77 between the FFQs delivered two weeks apart. These data demonstrate good agreement between our culturally tailored FFQ and 24DR, and moreover, robust reproducibility for quantifying intakes of food groups associated with hypertension in Nigeria. This confirms that this novel FFQ is a valid and reliable tool for assessing the intake of key food groups among Nigerian adults.

## Introduction

Hypertension is a leading risk factor for cardiovascular disease [1], which is annually attributes to over 10 million deaths worldwide [2, 3]. The highest hypertension burdens exist in low- and middle-income countries, with over 30% of adults affected in some African regions [4, 5]. In Nigeria specifically, hypertension prevalence has more than doubled since 1990, from 11.4% to 24.8% in 2015, with just over a quarter of hypertensive adults achieving blood pressure control [6, 7]. This escalating epidemic underscores an urgent need for improved screening, treatment, and prevention strategies.

Unhealthy dietary patterns are a predominant modifiable risk factor for hypertension globally, including in Sub-Saharan Africa [8, 9]. Specifically, diets characterised by elevated levels of saturated fat, processed meats, and sugar-sweetened beverages have been associated with an increased risk of hypertension [10]. Moreover, deficiencies in vital dietary components like fruits, vegetables, nuts, legumes, and omega-3 fatty acids from seafood correlate with elevated blood pressure levels [11]. In our prior research, we highlighted a significant association between high consumption of diets rich in dietary salt, red meat, processed foods, fried foods, dietary fat, and alcohol and an elevated risk of hypertension with a mean overall risk increase of 1.42 not only in Nigeria but also in various West African countries [12]. The average daily sodium intake in Nigeria ranges from 9-12 grams, which exceeds the World Health Organization’s (WHO’s) recommended limit of 5 grams [13]. This highlights dietary optimisation as a crucial component of population-level and clinical hypertension prevention strategies in Nigeria.

Dietary assessment represents an essential first step for establishing the association between diet and chronic diseases and designing effective public health dietary prevention strategies against chronic conditions, including hypertension [14]. To ensure optimal relevance and validity, dietary assessment tools must be customised to the cultural context of the population, encompassing region-specific foods, meals, serving sizes, and eating patterns [15]. Food frequency questionnaires (FFQs) developed for Western populations show limited validity for immigrant groups retaining traditional diets [16]. This underscores the need for culturally adapted FFQs, especially in ethnically diverse countries like Nigeria, which has over 250 ethnic groups [17]. However, culturally specific and validated FFQs for assessing dietary status, particularly for hypertension, are notably lacking in Nigeria and other West African countries, impeding progress in diet-disease research and clinical support.

In this study, following a robust validation protocol in a clinical setting in Nigeria, the validity of a novel culturally tailored FFQ was evaluated against three repeat 24-hour dietary recalls (24DR). Through assessing reproducibility, we sought to enhance the applicability of the FFQ in hypertension management and provide crucial insights for implementing the FFQ tailored to the Nigerian population.

## Materials and methods

### Study design and setting

This was a single-centre study incorporating both quantitative and qualitative methodologies. We sought to assess the relative validity and reproducibility of a newly developed tailored dietary screening tool consisting of a 27-food groups that we aim to incorporate into routine clinical practice in Nigeria to identify adults at high risk of hypertension. The investigation was conducted at the Internal Medicine and Family Medicine Department outpatient clinics of Rivers State University Teaching Hospital (RSUTH) in Port Harcourt, Rivers State, Nigeria.

### Ethics approval

The study protocol underwent review by two ethics boards. Firstly, it was submitted to the Business, Earth & Environment, Social Sciences (AREA FREC) Committee at the University of Leeds, Leeds, United Kingdom, on the 21st of March 2023. Subsequently, it was presented to the Rivers State University Teaching Hospital Research Ethics Committee in Port Harcourt, Nigeria, on March 20th, 2023. Final approvals were granted with the following reference numbers: 0484 on 28/04/2023 and RSUTH/REC/2023316 on 30/03/2023, respectively. The trial was duly registered at *clinicaltrials.gov* under the identifier NCT05973760.

### Eligible participants

Our study enrolled adult patients between the ages of 18 to 70 years attending the Rivers State University Teaching Hospital for their routine medical care, including both men and women, who had been residing in Nigeria for at least two years and possessed proficiency in reading, writing, and communicating in English. The complete list of inclusion and exclusion criteria are present in **Table 1**.

**Table 1:** Inclusion and exclusion criteria.

| Inclusion criteria | Exclusion criteria |
| --- | --- |
| Age between 18 and 70 years | Individuals < 18 years or > 70 years of age |
| Men and women | Pregnant women or intent to become pregnant or breastfeeding woman |
| Hypertensive or non-hypertensive individual | Diagnosis of other chronic diseases such as cancer, diabetes, renal failure, endocrine diseases, and previous and recent incidence of cardiovascular disease (CVD) and stroke |
| Individuals who have been residents in Nigeria for the past 2 years | Individuals who have been resident in Nigeria for shorter than 2 years |
| Ability to read, write, and communicate over the phone in English | Individuals on dietary restriction or recent changes to their diet or food |
| Individuals who gave their consent to participate | Individuals who did not give their consent to participate or are currently enrolled in other studies |

### Participant recruitment and informed consent

Participant recruitment occurred over four weeks in July 2023 during regular clinic visits. This process was facilitated through strategically placed recruitment posters within the hospital premises, referrals from healthcare professionals, and morning briefing sessions at the outpatient clinics of the Internal Medicine and Family Medicine Departments of RSUTH. Patients expressing interest in the study were screened for eligibility using a structured questionnaire (**Table 1**). Subsequently, eligible patients were categorised into either the hypertension or non-hypertension groups. The study adheres to SPIRIT guidelines for reporting clinical trials [18]. Before participation, each participant received and reviewed a simplified version of the participant information sheet. They had the opportunity to address any queries or concerns with the study personnel, ensuring their consent to participate was voluntary and fully informed. All patients provided written informed consent before participating in the study

### Sample size and sampling technique

Previous validation studies investigating the correlation between FFQs and 24DR have demonstrated good agreement, with correlation coefficients (*r*_s_) ranging from 0.3 to 0.7 [19–22]. A moderate *r*_s_ = 0.5 is typically considered a robust indicator of correlation [23]; therefore, it was used to estimate the sample size along with a statistical power of P = 0.8, a 95% confidence interval, and a two-tailed alpha level of 0.05 utilising the G*Power software [24]. While a minimum of 29 participants was determined to be required to accommodate an anticipated dropout rate of 20% and address any potential missing or incomplete data, we set the target sample size at 50 participants [25, 26]. Eligible participants were recruited through a non-probability convenience sampling method.

### Dietary Assessment

#### Dietary screening tool

The dietary screening tool was a newly developed, 27-food group semi-quantitative food frequency questionnaire (FFQ) that was designed to capture the usual food intake over the past month. The initial list of commonly consumed foods was created through guidance from the Nigerian and Ghana National Nutritional Guideline on Non-Communicable Disease Prevention, Control and Management [27, 28] and supported by our prior systematic review and meta-analysis the investigated dietary factors associated with hypertension in the West Africa [12]. Designed to be completed in less than 20 minutes, the final FFQ comprised 26 questions on major food groups and an additional 6 questions related to salt (**S1 Table**). These food groups encompassed various foods such as fruit, vegetable, fibre-breakfast cereals, rice and pasta, beans, yam and potatoes, fried or fast foods, whole meat, white meat, processed meat, sugary fizzy drinks and fruits, diet non-alcoholic drinks, tea and coffee, soups and stew (fatty soups, vegetable soups, draw soups, native soups, and stews), nuts and seeds, dessert and sweets, fats and oils, salt, milk and milk-based beverages (**S1 Table**). For each food item, participants were asked about the frequency of consumption over the past month, with response options ranging from ‘rarely or never,’ ‘1-2 times/week,’ ‘3-5 times/week,’ to ‘daily,’ and ‘more than once per day’ (**S1 Table**).

### 24-hour Dietary Recalls (24DR)

Three repeat 24-hour dietary recalls (24DR) were conducted by trained nutritionists on non-consecutive days, covering two weekdays and one weekend day. This approach aimed to account for the day-to-day variation in dietary intake. Throughout the recalls, detailed descriptions of all foods, snacks and beverages consumed in the preceding 24 hours were recorded, including cooking methods and brand names (where possible).

### Data Collection

Data collection was over four weeks in August 2023. At clinic visit 1 in week 1, the eligible consenting patients completed sociodemographic and health status questionnaires and underwent baseline assessments, including height, weight, and blood pressure measurement (**Fig 1**). The height and body weight were measured twice using a standard stadiometer (model number: DG2301, China), and the Body mass index (BMI) was calculated based on the measurements from the height and weight using the formula BMI= body weight in Kg/ (height in metre)^2^. The participant’s blood pressure was recorded twice in the non-dominant arm using an automated mercury sphygmomanometer (model number: ZK-BB68, Shenzhen, China).

**Fig 1.**
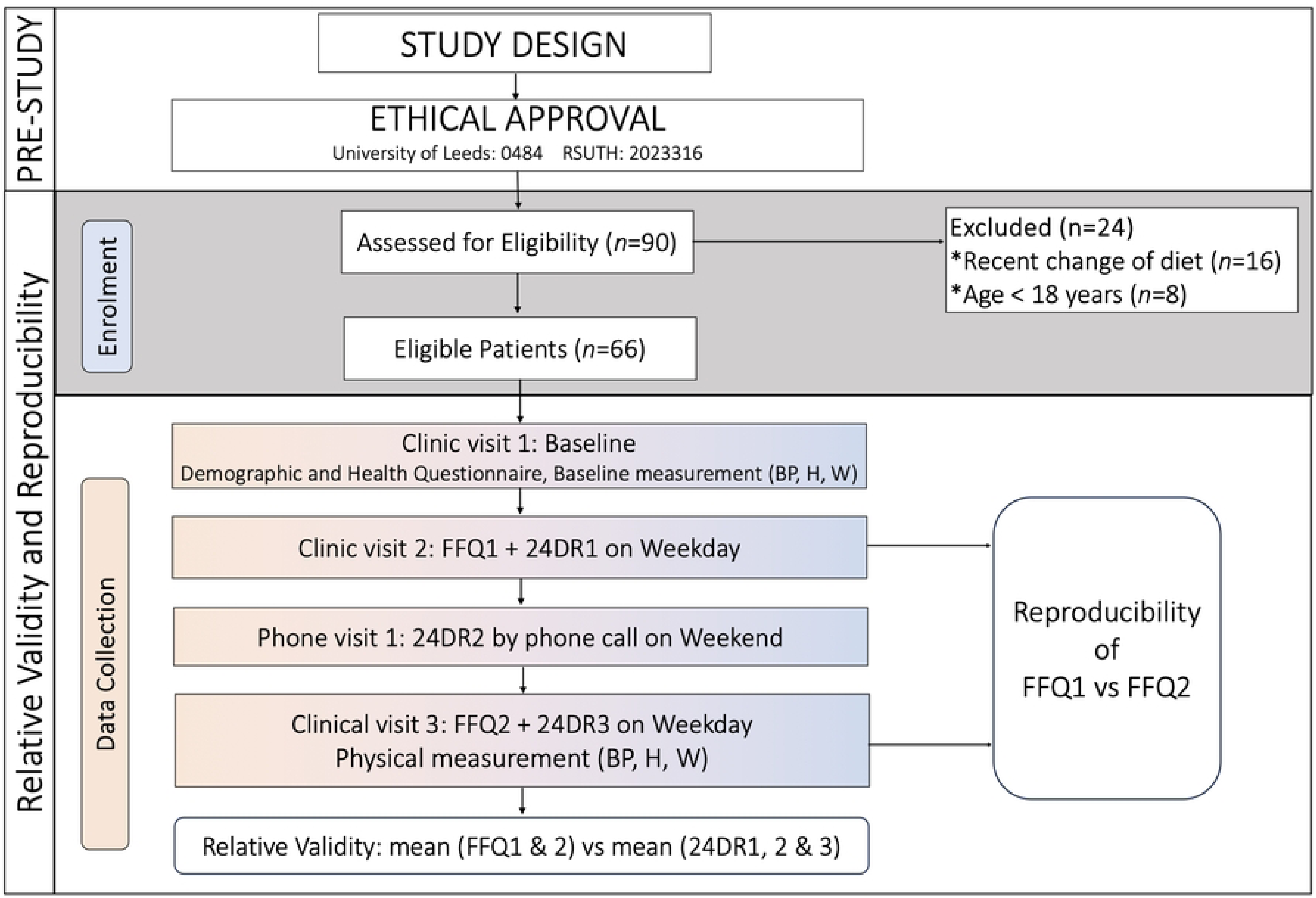
Study design, patient recruitment, enrolment, and data collection flowchart. FFQ1: first food frequency questionnaire, FFQ2: second food frequency questionnaire, 24DR1: first 24-hour dietary recall, 24DR2: second 24-hour dietary recall, 24DR3: third 24-hour dietary recall, BP: Blood pressure, H: Height, W: Weight.

At clinic visit 2 in week 2, eligible consenting patients completed the first self-administered dietary screening (FFQ1) tool. In conjunction with the FFQ1, the first interview-based 24DR (24DR1) was conducted to collect patient food intake data for the past 24 hours on one weekday using the multiple-pass method [29–31]. The second 24DR (24DR2) was conducted by phone to collect the patients’ food intake on a weekend day in week 2 (**Fig 1**). During clinic visit 3, the study patients completed the FFQ for the second time (FFQ2). The third 24DR (24DR3) was conducted on another weekday to collect food intake data for the past 24 hours. Finally, a second measurement of height, weight, and blood pressure was collected (**Fig 1**).

### Participant compensation

Upon completing all the requirements of the study protocol, consisting of the two FFQs and the three 24DR, and including all the physical measurements (weight, height, and blood pressure), patients were given a token of appreciation in the form of a £5 gift (equivalent to approximately ₦6,000). This was extended to participants as a gesture of gratitude for their participation and compensation for their valuable time.

### Statistical analysis

Dietary data (i.e., frequencies of food group intakes) from the first FFQ (FFQ1), second FFQ (FFQ2) and the three repeats 24-hour dietary recalls (24DR1, 24DR2 and 24DR3) for each participant were entered into a Microsoft Excel spreadsheet with quality checks. The frequencies of intakes reported in FFQ1 and FFQ2 were converted into servings/day by multiplying the average servings per week and then dividing the average by 7 days according to the method by Fatihal et al. [32]. For example, 3-5 times/ week was converted ([3+5/2] ÷ 7 days) into 0.57 serving/day. The salt intake assessed by the FFQ was coded numerically as ‘1 for ‘never or rarely’, 2 for ‘sometimes’, 3 for ‘usually’ and 4 for ‘always’. The food intake data from the FFQ and 24DR data were aggregated into 20 major food groups **(S2 Table).** The mean of the FFQ was calculated by combining the data from both administrations (FFQ1 and FFQ2). Additionally, the mean for the 24DR was computed based on three non-consecutive repetitions of the 24DR. These means were used for the validity analysis.

The frequency data and the mean differences between the FFQ and 24DR were tested for normality using the Shapiro-Wilk [33] and Kolmogorov-Smirnov tests [34] with inspection of the histogram. The data were not normally distributed; therefore, non-parametric methods were used for the analysis. The results were reported as mean, median and interquartile range (IQR) for continuous data and n (%) for categorical data. P-values <0.05 were considered statistically significant. All analyses were performed using R Statistical Software (v4.3.1) [35]. The statistical analyses were performed in 2 phases.

In the first phase, we used multiple methods to assess the relative validity of the FFQ by evaluating the agreement between the mean of the FFQ and the mean of the 24DR. First, Spearman’s rank correlation was used to compare the frequency of food group intakes from the FFQ with those from the 24DR. A correlation coefficient above 0.5 indicated a good correlation [36]. Secondly, the Wilcoxon signed-rank test compared the difference between the mean FFQ and mean 24DR for each food group. A *p*-value > 0.05 was considered to indicate no statistically significant difference and good agreement [37, 38]. Thirdly, the cross-classification of intakes into quartiles by the 3 methods (FFQ and 24DR): (i) proportion of exact agreement, deviation by 1 quartile; (ii) proportion of adjacent agreement, indicating deviation by adjacent quartiles; and (ii) proportion of grossly misclassified participants, disagreement by 3 quartiles. Finally, the Bland-Altman analysis [39] was used to assess the level of agreement and whether differences between FFQ and 24DR estimated measurements were dependent on the magnitude of measurements. The mean difference (mean FFQ– mean 24DR) was plotted against the average of the two measures ([mean FFQ + mean 24DR)/2]) for each food group. An acceptable level of agreement was defined as differences in means falling within the range of ±3 standard deviations (SDs) [40]. Additionally, the relative differences (%) within this range were calculated to quantify agreement.

In phase 2, we assessed the reproducibility of the FFQ at two different administrations (FFQ1 vs FFQ2). The strength and association of the FFQ1 and FFQ2 were evaluated using Spearman’s rank correlations. The agreement and consistency between food groups from the two FFQ administrations were assessed using intraclass correlation coefficients (ICCs). ICC values were calculated based on a single rating, absolute agreement, and 2-way mixed-effects model [41]. ICC values above 0.60 were considered evidence of good reproducibility between the two FFQ administrations [36]. The ranking agreement between the FFQ1 and FFQ2 was evaluated using the Wilcoxon signed-rank test, and a *p*-value >0.05 was considered to indicate a good agreement between the FFQ1 and FFQ2.

## Results

### Participant characteristics

A total of 90 patients indicated interest in the study. Of these, 66 met the inclusion criteria and consented to participate in the study. Of the 66 eligible consenting patients, 58 completed the study protocol and their data were included in the final data analysis (**Fig 2**). The overall average age was 42.6 ± 11.9 years, with hypertensive participants being older, on average 46.4 ± 10.1 years, compared to non-hypertensives with a mean age of 38.7 ± 12.4 years. The majority of participants were female (69%) and over two-thirds (69%) had university or postgraduate education. Family history of hypertension was reported by 55.2% (**Table 2**).

**Fig 2.**
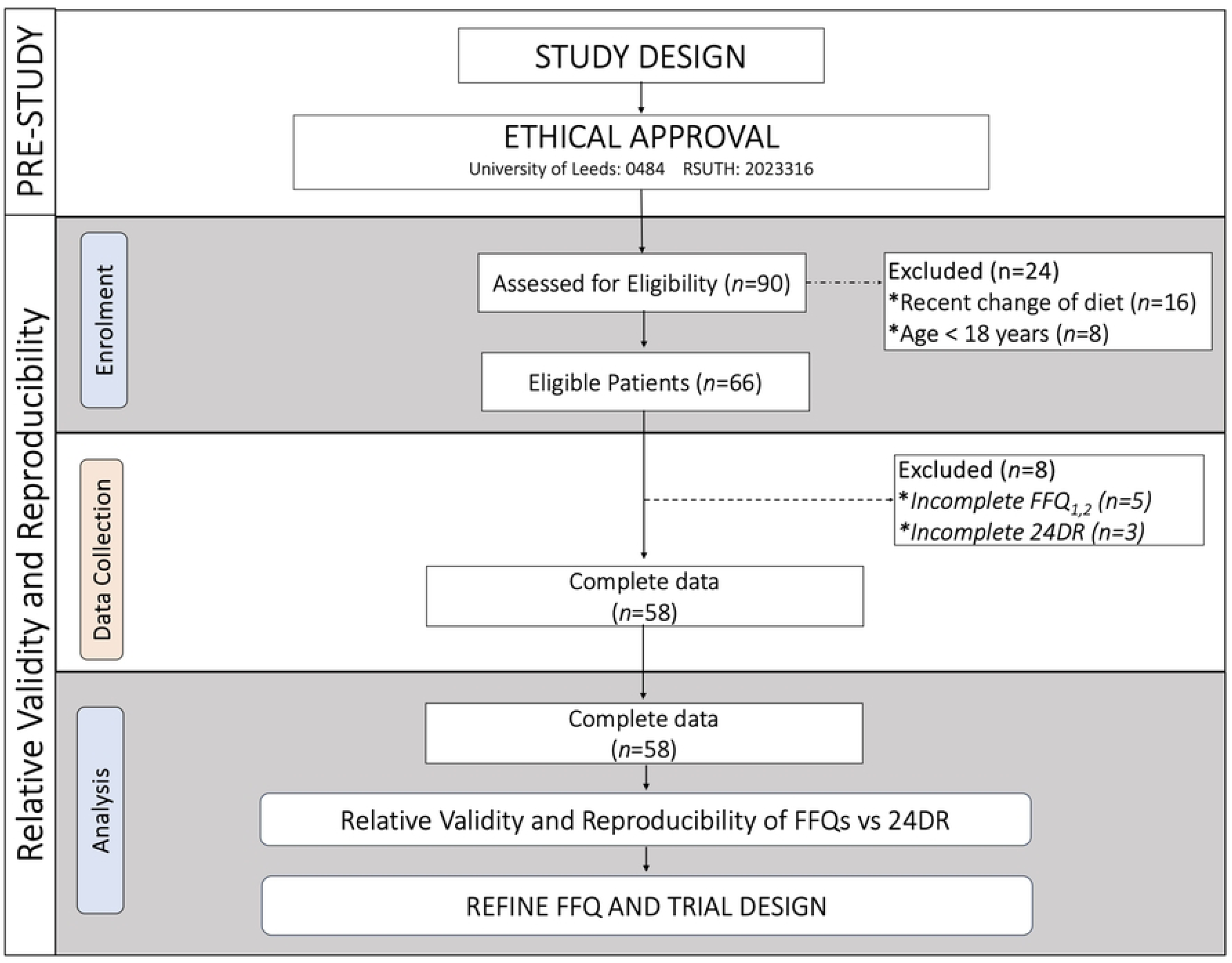
Participant selection and sequence of assessments flowchart. FFQ: food frequency questionnaire, 24DR: 24-hour dietary recalls.

**Table 2.** Sociodemographic, anthropometric, and clinical characteristics of participants.

| Characteristics | Overall (n=58) | Non-hypertensive (n=29) | Hypertensive(n=29) |
| --- | --- | --- | --- |
| Age (years), mean ± SD | $42.6 \pm 11.9$ | $38.7 \pm 12.4$ | $46.4 \pm 10.1$ |
| Sex, n (%) |  |  |  |
| Male | 18 (31.0) | 9 (31.0) | 9 (31.0) |
| Female | 40 (69.0) | 20 (69.0) | 20 (69.0) |
| Education n (%) |  |  |  |
| Primary | 2 (3.5) | 1 (3.5) | 1 (3.5) |
| Secondary | 12 (20.7) | 3 (10.3) | 9 (31.0) |
| High school | 4 (6.9) | 2 (6.9) | 2 (6.9) |
| University | 26 (44.8) | 14 (48.3) | 12 (41.4) |
| Postgraduate | 14 (24.1) | 9 (31.0) | 5 (17.2) |
| Family history of Hypertension, *n* (%) |  |  |  |
| Yes | 32 (55.2) | 19 (65.5) | 13 (44.8) |
| Years of hypertension *n* (%) |  |  |  |
| < 1 year | 9 (31.0) | NA | 9 (31.0) |
| 1-5 years | 8 (27.6) | NA | 8 (27.6) |
| > 5 years | 12 (41.4) | NA | 12 (41.4) |
| Antihypertensive medications use *n* (%) |  |  |  |
| Yes | 16 (55.2) | None | 16 (55.2) |
| No | 13 (44.8) | None | 13 (44.8) |
| Height (m) | 1.65 ± 0.1 | 1.68 ± 0.1 | 1.62 ± 0.1 |
| Body weight (kg) | 79.4 ± 17.2 | 75.0 ± 15.4 | 83.8 ± 18.1 |
| Body mass index (kg/m<sup>2</sup>) | 29.5 ± 7.1 | 26.9 ± 6.8 | 32.1 ± 6.4 |
| Blood pressure |  |  |  |
| SBP (mmHg) | 140.3 ± 23.9 | 121.0 ± 11.7 | 159.0 ± 16.9 |
| DBP (mmHg) | 87.4 ± 17.3 | 75.4 ± 9.7 | 99.3 ± 14.8 |

Data are presented as: *n*= frequency, %= percentage, mean ± SD= Standard deviation, NA = not applicable

A considerable proportion of participants had experienced hypertension for more than 5 years (41.4%), but only 55.2% reported using antihypertensive medications. Participants with hypertension, on average, appeared to be heavier (83.8kg vs 75kg), with more presenting with obesity (mean+/- SD BMI: 32.1 ± 6.4 kg/m^2^) than those who did not have hypertension (26.9 ± 6.8 kg/m^2^). Similarly, participants with hypertension, on average, had higher systolic blood pressures (159.0 ± 16.9 mmHg vs 121.0 ± 11.7 mmHg) in spite of a high percentage using antihypertensive medications (**Table 2**).

### Dietary intake assessment

The mean and median servings/day were similar between the two dietary assessment methods for most food groups (**Table 3**). The mean fold differences varied from 0.25 for fats and oils to 1.25 for yam and potatoes, indicating that, on average, the FFQ provided food groups intake estimates within 75% below to 25% above the 24DR amounts. Overall, no significant differences were observed between the food group intakes estimated by the FFQ and the 24DR.

**Table 3.** Food group intake estimates from the novel food frequency questionnaire (FFQ) and the 24-hour dietary recall (24DR).

| Food Group<br>(servings/day) | FFQ |  |  | 24DR |  |  | Mean Fold Difference<br>(FFQ/24DR) |  |  |
| --- | --- | --- | --- | --- | --- | --- | --- | --- | --- |
|  | Mean | Median | IQR | Mean | Median | IQR | Mean | 95% CI | p-value |
| Fruit | 0.38 | 0.21 | 0.57 | 0.35 | 0.33 | 0.67 | 1.09 | 0.82; 1.41 | 0.288 |
| Vegetables | 0.48 | 0.21 | 0.36 | 0.46 | 0.39 | 0.36 | 1.05 | 0.94; 1.19 | 0.193 |
| Grains | 0.40 | 0.33 | 0.19 | 0.42 | 0.44 | 0.20 | 0.96 | 0.84; 1.10 | 0.741 |
| Beans and lentils | 0.33 | 0.21 | 0.36 | 0.34 | 0.33 | 0.26 | 0.99 | 0.82; 1.12 | 0.723 |
| Meat | 0.55 | 0.50 | 0.30 | 0.61 | 0.50 | 0.29 | 0.90 | 0.80; 1.00 | 0.975 |
| Processed meat | 0.16 | 0.00 | 0.21 | 0.14 | 0.00 | 0.00 | 1.19 | 0.95; 1.59 | 0.069 |
| Fish and seafoods | 0.62 | 0.39 | 0.58 | 0.54 | 0.33 | 0.59 | 1.16 | 0.96; 1.36 | 0.058 |
| Eggs | 0.32 | 0.21 | 0.36 | 0.31 | 0.33 | 0.67 | 1.04 | 0.85; 1.28 | 0.365 |
| Fried or fast food | 0.23 | 0.21 | 0.37 | 0.29 | 0.33 | 0.33 | 0.80 | 0.62; 1.03 | 0.954 |
| Yam and potatoes | 0.30 | 0.21 | 0.18 | 0.25 | 0.33 | 0.33 | 1.25 | 0.88; 1.85 | 0.136 |
| Soups | 0.35 | 0.32 | 0.22 | 0.33 | 0.33 | 0.23 | 1.08 | 0.96; 1.20 | 0.096 |
| Stew | 0.45 | 0.39 | 0.36 | 0.42 | 0.33 | 0.34 | 1.09 | 0.92; 1.27 | 0.164 |
| Nuts and seeds | 0.46 | 0.29 | 0.36 | 0.44 | 0.33 | 0.67 | 1.06 | 0.86; 1.29 | 0.298 |
| Desserts and sweets | 0.19 | 0.00 | 0.21 | 0.21 | 0.00 | 0.33 | 0.96 | 0.60; 1.48 | 0.615 |
| Soft drinks | 0.22 | 0.13 | 0.24 | 0.19 | 0.17 | 0.33 | 1.15 | 0.85; 1.57 | 0.222 |
| Alcoholic drinks | 0.03 | 0.00 | 0.00 | 0.04 | 0.00 | 0.00 | 1.00 | 0.43; 1.57 | 0.787 |
| Tea and coffee | 0.39 | 0.21 | 0.47 | 0.34 | 0.33 | 0.67 | 1.15 | 0.89; 1.47 | 0.153 |
| Milk and milk drinks | 0.46 | 0.39 | 0.36 | 0.48 | 0.33 | 0.34 | 0.96 | 0.82; 1.11 | 0.739 |
| Fats and oils | 0.58 | 0.57 | 0.74 | 2.30 | 2.33 | 0.92 | 0.25 | 0.20; 0.31 | 1.000 |
| Salt and seasonings | 3.37 | 4.00 | 1.00 | 3.45 | 3.33 | 1.33 | 0.98 | 0.88; 1.08 | 0.672 |

FFQ: Food frequency questionnaire; 24DR: 24-hour dietary recalls; IQR: Interquartile range

### Assessment of relative validity

To assess the validity of the FFQ, we evaluated the relationship between the food group intake estimated by the FFQ relative to the 24DR. The Spearman’s correlation coefficients (*r*_s_) ranged from 0.20 for fats and oils to 0.78 for vegetables, with an average **c**orrelation coefficient of 0.60 (**Table 4**). Although weaker positive correlation coefficients (*r*_s_<0.3) were found for fat & oils, and salt, most of the food groups (*n*=15) had a correlation coefficient ≥ 0.50, indicating a strong positive correlation between the mean FFQ and mean 24DR (*p*<0.05). In addition, among the 20 food groups evaluated in the FFQ, 19 food groups had no significant differences (*p*>0.05) in the mean and median intakes compared to 24DR when a Wilcoxon signed-rank test was applied — the exception was ‘fats and oils’ (*p*<0.05) (**Table 4**). Overall, the results suggest that the FFQ provides comparable rankings and intake estimates for most foods (*n*=19) compared to 24DR and shows good agreement between the dietary assessment approaches.

**Table 4.** Comparison of food group intake obtained from a semi-quantitative food frequency questionnaire (FFQ) and the 24-hour dietary recalls (24DRs).

| Food Group<br>(servings/day) | Agreement between FFQ and 24DR |  |  | Disagreement between FFQ and 24DR |  |  |
| --- | --- | --- | --- | --- | --- | --- |
| | $r_s$ | <sup>a</sup>$p$-value | <sup>b</sup>$p$-value | Exact (%) | Adjacent (%) | GM<sup>1</sup>(%) |
| Fruit | 0.65 | <0.001 | 0.748 | 53 | 33 | 14 |
| Vegetables | 0.78 | <0.001 | 0.706 | 50 | 45 | 5 |
| Grains | 0.64 | <0.001 | 0.042 | 40 | 53 | 7 |
| Beans and lentils | 0.64 | <0.001 | 0.632 | 53 | 40 | 7 |
| Meat | 0.65 | <0.001 | 0.063 | 50 | 43 | 7 |
| Processed meat | 0.74 | <0.001 | 0.215 | 88 | 10 | 2 |
| Fish and seafoods | 0.72 | <0.001 | 0.869 | 62 | 35 | 3 |
| Eggs | 0.77 | <0.001 | 0.224 | 28 | 55 | 17 |
| Fried or fast food | 0.48 | <0.001 | 0.081 | 52 | 33 | 15 |
| Yam and potatoes | 0.37 | 0.004 | 0.619 | 45 | 45 | 10 |
| Soups | 0.66 | <0.001 | 0.361 | 48 | 45 | 7 |
| Stew | 0.62 | <0.001 | 0.174 | 47 | 47 | 7 |
| Nuts and seeds | 0.71 | <0.001 | 0.862 | 66 | 31 | 3 |
| Desserts and sweets | 0.47 | <0.001 | 0.237 | 64 | 26 | 10 |
| Soft drinks | 0.65 | <0.001 | 0.806 | 69 | 29 | 2 |
| Alcoholic drinks | 0.63 | <0.001 | 0.287 | 88 | 12 | 0 |
| Tea and coffee | 0.55 | <0.001 | 0.501 | 48 | 38 | 14 |
| Milk & milk drinks | 0.75 | <0.001 | 0.338 | 67 | 26 | 7 |
| Fats and oils | 0.20 | 0.135 | <0.001 | 22 | 59 | 19 |
| Salt and seasonings | 0.22 | 0.154 | 0.968 | 17 | 24 | 59 |

<sup>1</sup> Gross misclassification, disagreement by 3 quartiles, $r_s$ : Spearman's correlation coefficient, <sup>a</sup>: $p$ -value of Spearman's
Rank correlation coefficients, <sup>b</sup> $p$ -value: $p$ -value for Wilcoxon signed-rank test of difference. FFQ: Food frequency questionnaire; 24DR: 24-hour dietary recalls

Additionally, the percentage of participants grossly misclassified by 3 quartiles ranged from 0% for alcoholic drinks to 59% for salt, with an average of 11% (**Table 4**). For most food groups (*n*=15), over 50% of the participants were classified into the same or neighbouring quartile. Specifically, the classification of participants into the exact or adjacent quartiles ranges from 10% for dessert and sweets to 88% for processed meat and alcoholic drinks, with an average exact agreement of 53% and an adjacent agreement of 37% (**Table 4**). Importantly, 90% of participants were classified in the same or neighbouring quartile, indicating a good agreement between the FFQ and 24DR.

Furthermore, the Bland-Altman analysis was used to assess the level of agreement between the FFQ and 24DR (**S3 Table**). **Fig 3A-F** presents the Bland-Altman plots for the 3 healthy food groups of the DASH diet (e.g., fruits, vegetables, and nuts and seeds) [42] and 3 less healthy food groups/items identified by our recent meta-analysis of foods associated with hypertension in West African countries, including Nigeria [12] (e.g., salt, fried/fast foods, and fats and oils). The plots for the remaining food groups are provided in supplementary materials (**S1 Fig).** Although moderate bias and wide limits of agreement (-4.18 to 3.93) were observed for fats/oils and salt food groups (**Fig 3D-F**), very limited bias was observed for the majority (*n*=18) of the food groups, where mean differences (bias) ranged from -0.06 servings/day (meat and fried and fast foods) to 0.08 servings/day (fish) (**Fig 3A-3C and S1 Fig**). In addition, the 95% limits of agreement (LOA) spanned from -1.23 to -0.20 servings/day (lower LOA) to 0.19 to 1.40 servings/day (upper LOA) for most food groups (*n*=18), suggesting reasonable agreement (**S3 Table**). A high proportion (>96%) of observations fell within the acceptable limits of agreement (± 3 standard deviation LOA) without increased differences across higher food intake ranges **(S3 Table).** In summary, the Bland-Altman analysis and plots suggest a high level of agreement between the FFQ and 24DR for the majority of food groups assessed (*n*=18).

**Fig 3.**
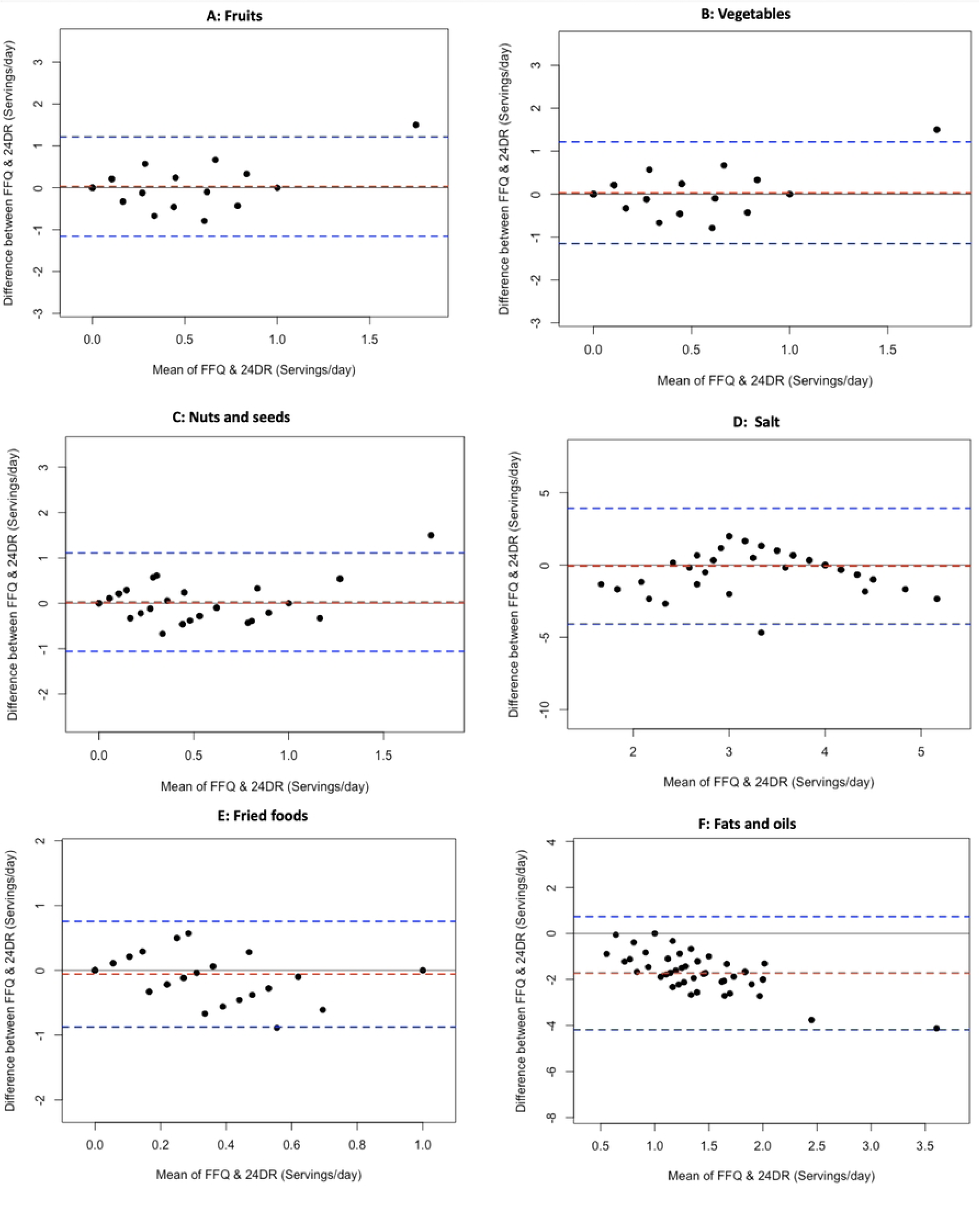
Bland-Altman plots related to food groups identified in the DASH diet (A: Fruit, B: Vegetables, C: Nuts and seeds) and D: Salts, E: Fried and fast foods and F: Fats and oils. Differences in the serving/day of food groups derived from the mean of the three repeat 24-hour recalls (24DR) and the mean of the food frequency questionnaire (FFQ) were plotted against the corresponding mean serving/day derived from the two methods. Dashed red lines represent the mean difference (bias), and dashed blue lines show the upper and lower 95% limits of agreement (*n*=58).

### Assessment of reproducibility

Assessing reproducibility between the two administrations of the FFQ, Spearman’s ranked correlation coefficient ranged from 0.38 for yam and potatoes to 0.97 for salt, with an average correlation coefficient of 0.75, with most food groups (17/20) showing correlation coefficients above 0.60. All correlation coefficients were statistically significant (*p*<0.001), reaffirming the high level of agreement between the two FFQs (**Table 5**). Additionally, among the 20 food groups evaluated for reproducibility, no significant differences in the mean and median intakes between the FFQ1 and FFQ2 were observed in the Wilcoxon signed-rank test for all the food groups (*p*>0.05) (**Table 5**).

**Table 5.** Reproducibility on the number of food group serving/day estimated by repeated administration of a Food Frequency Questionnaire.

| Food Groups | FFQ1 | FFQ2 | Reproducibility (FFQ1 and FFQ2) |
| --- | --- | --- | --- |

| (Servings/day) | Mean | Median | IQR | Mean | Median | IQR | $r_s$ | *$p$-value | ICC | 95%CI |
| --- | --- | --- | --- | --- | --- | --- | --- | --- | --- | --- |
| Fruit | 0.38 | 0.21 | 0.57 | 0.39 | 0.21 | 0.57 | 0.90 | 0.750 | 0.98 | 0.96; 0.98 |
| Vegetables | 0.49 | 0.21 | 0.36 | 0.46 | 0.21 | 0.36 | 0.84 | 0.154 | 0.92 | 0.88; 0.95 |
| Grains | 0.43 | 0.21 | 0.57 | 0.42 | 0.21 | 0.57 | 0.70 | 0.577 | 0.72 | 0.64; 0.79 |
| Beans and lentils | 0.32 | 0.21 | 0.36 | 0.33 | 0.21 | 0.36 | 0.91 | 0.479 | 0.87 | 0.80; 0.92 |
| Meat | 0.59 | 0.57 | 0.79 | 0.56 | 0.57 | 0.79 | 0.83 | 0.123 | 0.85 | 0.80; 0.90 |
| Processed meat | 0.16 | 0.00 | 0.21 | 0.15 | 0.00 | 0.21 | 0.79 | 0.751 | 0.87 | 0.79; 0.92 |
| Fish and seafoods | 0.65 | 0.39 | 0.79 | 0.58 | 0.39 | 0.79 | 0.79 | 0.166 | 0.80 | 0.68; 0.88 |
| Eggs | 0.31 | 0.21 | 0.36 | 0.33 | 0.21 | 0.36 | 0.85 | 0.590 | 0.79 | 0.66; 0.87 |
| Fried or fast foods | 0.26 | 0.21 | 0.48 | 0.22 | 0.21 | 0.21 | 0.76 | 0.203 | 0.73 | 0.59; 0.83 |
| Yam and potatoes | 0.29 | 0.21 | 0.27 | 0.32 | 0.21 | 0.00 | 0.38 | 0.886 | 0.55 | 0.34; 0.71 |
| Soups | 0.35 | 0.21 | 0.36 | 0.21 | 0.21 | 0.36 | 0.65 | 0.783 | 0.75 | 0.69; 0.80 |
| Stew | 0.45 | 0.39 | 0.36 | 0.46 | 0.21 | 0.36 | 0.53 | 0.867 | 0.31 | 0.06; 0.53 |
| Nuts and seeds | 0.54 | 0.21 | 0.36 | 0.39 | 0.21 | 0.36 | 0.68 | 0.099 | 0.50 | 0.28; 0.67 |
| Desserts & sweets | 0.19 | 0.00 | 0.21 | 0.19 | 0.00 | 0.21 | 0.55 | 0.746 | 0.45 | 0.21; 0.63 |
| Soft drinks | 0.23 | 0.00 | 0.21 | 0.21 | 0.00 | 0.21 | 0.87 | 0.290 | 0.92 | 0.89; 0.95 |
| Alcoholic drinks | 0.04 | 0.00 | 0.00 | 0.03 | 0.00 | 0.00 | 0.71 | 0.773 | 0.94 | 0.91; 0.97 |
| Tea and coffee | 0.38 | 0.21 | 0.36 | 0.39 | 0.21 | 0.57 | 0.73 | 0.944 | 0.73 | 0.58; 0.83 |
| Milk & milk drinks | 0.44 | 0.21 | 0.36 | 0.46 | 0.39 | 0.36 | 0.92 | 1.000 | 0.95 | 0.92; 0.97 |
| Fats and oils | 0.57 | 0.57 | 0.79 | 0.57 | 0.57 | 0.79 | 0.62 | 0.985 | 0.72 | 0.57; 0.82 |
| Salt intake | 0.83 | 1.00 | 0.29 | 0.81 | 1.00 | 0.29 | 0.97 | 0.371 | 0.96 | 0.94; 0.98 |

FFQ1: First food frequency questionnaire administration, FFQ2: Second food frequency questionnaire administration, ICC: Intraclass correlation coefficients ( $p < 0.05$ ), CI= Confidence interval of ICC, $r_s$ : Spearman's rank correlation coefficient, IQR: Interquartile range, \* $p$ -value for the test of the difference between FFQ1 and FFQ2 by Wilcoxon signed-rank test.

Furthermore, the intraclass correlation coefficient (ICC) was used to evaluate the consistency and agreement between the FFQ1 and FFQ2 (**Table 5**). ICCs ranged from 0.31 for stew to 0.98 for fruit, with an average intraclass correlation coefficient of 0.77. The majority of food groups (*n*=17) had ICC ≥0.70, which, according to the criteria of Koo and Li [41] and Cade et al. [36], indicates good to excellent reproducibility (**Table 5**). These findings suggest good reproducibility and consistency in individual rankings and negligible between time points for the FFQ, confirming the test-retest reliability of the FFQ across the food groups evaluated.

## Discussion

This study is the first to test and validate a culturally sensitive food frequency questionnaire (FFQ) for dietary screening of men and women for high-risk dietary habits associated with hypertension in a Nigerian clinic. Our aim is for the tool to be used by clinicians and patients across West Africa to: (i) facilitate discussions of dietary habits and cardiovascular health in the clinical setting; (ii) inform personalised dietary advice for patients at risk or with hypertension; and (iii) empower its citizens to take an active role in managing hypertension. With the participation of 58 men and women, the FFQ demonstrated good validity and reproducibility in a clinical setting for assessing intakes of food associated with hypertension in Nigeria, compared to 24DR.

### Relative validation

In all validation studies, some under- and over-estimation is expected but must be within an acceptable range. Streppel et al. evaluated the validity of an FFQ against the 24DR among 128 Dutch adults and reported overestimation in 13 of 21 foods by the FFQ [43], while Steinemann et al. reported overestimation in 13 of 25 foods compared to a 4-day weighed food record among 56 participants in Germany and a correlation coefficients from 0.09 (soup) to 0.92 (alcohol) with 16 out of the 25 food groups having correlation coefficients <0.50 [6][21].

Our FFQ demonstrated similar measures of overestimation from 4-25% and correlation coefficients between the FFQ and 24DR from 0.20 to 0.78, with the majority of the food groups (n=15) demonstrating moderate to strong positive correlations (*r*_s_ >0.50), indicating good agreement.

Additionally, our study reported that 90% of participants were classified into the same or neighbouring quartiles when comparing FFQ and 24DR, where the exact agreement for most food groups (17/27) was 30-88% with an average gross misclassification of 11% across food groups. This is in agreement with other successful validation studies in Australia (n=96 adults) that reported 27-70% exact agreement and under 15% gross misclassification for most food groups [44, 45].

Finally, the Bland-Altman method [39] was used to illustrate the agreement between the FFQ and 24DR. Although ‘Fats and oils’ and salt were underestimated by the FFQ, as noted in other studies [46], the majority of the food groups assessed in our study demonstrated minimal bias. Indeed, > 96% of observations were within acceptable limits of agreement. This aligns with or exceeds the results of previous work, where FFQ validations study among (i)

130 men with prostate cancer reported similar small mean differences and acceptable agreements across 11 food groups [40]; and, (ii) 114 Lebanese adults with >80% agreement for the majority of the food groups [47]. Collectively, these results suggest that the validity of our novel FFQ meets or exceeds the levels of agreement reported by other validation studies and indicates that our tailored FFQ is well-designed for capturing the dietary habits of men and women in Nigerian populations.

### Reproducibility

The reproducibility of an FFQ is an important attribute to minimise bias [48]. The FFQ exhibited commendable reproducibility between the two collection points (FFQ1 vs FFQ2), yielding a robust positive correlation that surpassed *r*_s_ > 0.50 for the majority of assessed food groups and intraclass correlation coefficient > 0.70 for all the food groups assessed. These findings align with the results of other FFQ reproducibility studies that reported correlation coefficients between 0.32 and 0.87 [40] and 0.65 to 0.98 [47, 49] and agree with current recommended standards for reproducibility between 0.5 and 0.70 [36, 41, 50]. According to these criteria, our novel FFQ demonstrated good levels of agreement between baseline and follow-up. This suggests that it is well suited for accurately collecting dietary information and capturing dietary inconsistencies in Nigeria for clinicians [48], researchers, and public health professionals [51].

### Practical application and clinical relevance

The validity and reproducibility of our study data provide compelling evidence to further investigate the implementation and use of our novel FFQ as a practical clinical tool to screen and evaluate patient-mediated dietary risk for hypertension. The FFQ was able to rank intakes in food groups associated with hypertension [11, 12], including fruits, vegetables, grains, dairy, salt and fats and oil-based foods (soups and stew), to a similar degree of accuracy as 24DR, but was able to be completed in < 20 minutes. The results indicate that the FFQ can identify patients with high-risk dietary patterns who may benefit from prioritised education and support around dietary modification for blood pressure management. Integrating this rapid dietary screening tool into primary and tertiary care workflows will be a key step to enable a systematic approach to dietary monitoring and counselling in a clinical setting for hypertension prevention and management [52].

### Strengths and limitations

Although the study results underline the strength and potential of the FFQ in a clinical setting, we openly acknowledge some limitations: (i) 24DR has inherent limitations, including possible recall biases and within-person variability in daily intake, which can attenuate validation study results [53]. However, we aimed to reduce this by having a designated professional perform all 24DR evaluations and the use of repeat recalls on non-consecutive days, including two weekdays and one weekend day, which also mitigates this intra-individual variation; (ii) the use of a non-random, convenience sample may restrict the generalizability of findings to the broader target population; however, as this tool will be used in a clinical setting we foresee this limitation as minor and inherent to the tools primary purpose; (iii) literacy barriers among some participants required interviews instead of self-administration, which could have impacted their responses in the form of respondent bias/social desirability; and finally (iv) testing the FFQ in a relatively small geographic area reduces the applicability of results more widely across diverse Nigerian populations, which is something we aim to address in future work.

Nonetheless, despite these limitations, this study had several important strengths: (i) the use of multiple 24DR as the reference method provided detailed participant-informed dietary intake data and allowed for day-to-day variability to be assessed, thereby strengthening the quality of the reference data; (ii) the use of multiple analytical methods permitted a comprehensive assessment of the agreement between the FFQ and the 24DR; (iii) evaluation of the reproducibility or test-retest reliability of the FFQ two weeks apart permitted the reliability and consistency of the FFQ to be estimated over time, which will offer a better measure of habitual dietary habits; and finally (iv) testing the FFQ in the patient demographic, clinical setting and cultural context of its intended use improves the applicability of the validation.

## Conclusion

This study provides important evidence that the culturally tailored FFQ has adequate relative validity and reproducibility for ranking dietary intake of major foods and food groups in a clinical setting, compared to the average of three repeat non-consecutive 24DR. Therefore, we offer a valid short FFQ that could help assess common food group intakes for the assessment and prevention of hypertension, which could empower clinicians, patients, and researchers to take an active role in preventing hypertension in Nigeria and other West African countries. Further refinements of the FFQ and future studies on the acceptability of the FFQ will improve validity for some food groups.

## Data Availability

The values behind the means, standard deviations and other measures are reported and the data has been made privately available for reviewers at: https://data.4tu.nl/private_datasets/LG8jdbsGJQz5hA6vL1ik_-5OvufCdpcAeHnIdtbP-n8. We have embargoed the data for release until publication of the manuscript or the lead author (PhD student) has completed their PhD (August 2024), whoever happens first.

## Acknowledgement

We would like to extend our heartfelt gratitude to the patients and medical professionals at Rivers State University Teaching Hospital in Port Harcourt, Nigeria. Their invaluable participation and feedback were critical for the successful implementation of this research. In particular, we are thankful to Drs. Nkiru Ahiakwo, Ifeoma Enyoghasim, Eneyo Nelly, Comfort Imarhiagbe, Janny Ikurayeke, Valentine Kogbara, Titi Owen, Dickson Christian, Anwuri Grend, Anita Oweredaba, Ununuma Oguzor and Josephine Sokolo from the Family Medicine Department for their qualitative inputs on refining the food frequency questionnaire screening tool. We also sincerely appreciate the insights provided by Drs. Ibieneiyi, Edith Reuben, Elile Okpara, Imaobong Nonju, Siya George-Batubo, Chinnasa Nzokurum and Prof. Amah-Tariah, which significantly enhanced the quality of our study. In addition, we are grateful to the hospital management and the Internal Medicine and Family Medicine Departments for granting us ethical approval and access to facilities, which facilitated the seamless execution of this project.

## Authors’ Contributions

NPB and MAZ collaborated on the research methodology design. NPB led the trial. NPB led the analysis and prepared the first draft. MAZ and JBM provided analytical expertise. MAZ, JBM, and CIA provided critical feedback. NPB revised the manuscript. All authors approved the final manuscript.

## Funding

This study was funded by the Tertiary Education Trust Fund (TETFund) of Nigeria. MAZ is currently funded by Wellcome Trust (217446/Z/19/Z). The funders do not have any role in any aspect of this study. The funders had no role in study design, data collection and analysis, decision to publish, or preparation of the manuscript.

## Conflicts of Interest

The authors have declared that no competing interests exist.

## Supporting information

**S1 Table:** Food Frequency Questionnaire

**S2 Table**: Categorisations of food groups in the food frequency questionnaire (FFQ) corresponding to the FFQ question numbers.

**S3 Table:** Assessment of agreement between the food intake estimation of the food frequency questionnaire (FFQ) and 24-hour dietary recalls (24DR) using Bland-Altman analysis.

**S1 Fig.** Bland-Altman plots related to food groups.

## Notes

### Competing Interest Statement

The authors have declared no competing interest.

### Clinical Trial

NCT05973760

### Clinical Protocols

https://www.medrxiv.org/content/10.1101/2023.09.25.23296109v1

### Author Declarations

The study protocol underwent review by two ethics boards. Firstly, it was submitted to the Business, Earth & Environment, Social Sciences (AREA FREC) Committee at the University of Leeds, Leeds, United Kingdom, and was granted ethical approval (0484) on 28/04/2023. Secondly, it was presented to the Rivers State University Teaching Hospital Research Ethics Committee in Port Harcourt, Nigeria, and received ethical approval (RSUTH/REC/2023316) on 30/03/2023.

## References

1. Forouzanfar MH, Liu P, Roth GA, Ng M, Biryukov S, Marczak L, et al. Global Burden of Hypertension and Systolic Blood Pressure of at Least 110 to 115 mm Hg, 1990-2015. JAMA. 2017;317(2):165–82 DOI: 10.1001/jama.2016.19043.

2. GBD. Risk Factors Collaborators. Global burden of 87 risk factors in 204 countries and territories, 1990-2019: a systematic analysis for the Global Burden of Disease Study 2019. Lancet. 2020;396(10258):1223–49 DOI: 10.1016/S0140-6736(20)30752-2.

3. Saloni D, Fiona S, Hannah R, Max R. “Deaths by risk factor” Published online at OurWorldInData.org. 2023, ‘https://ourworldindata.org/causes-of-death<u>’</u>

4. Mills KT, Bundy JD, Kelly TN, Reed JE, Kearney PM, Reynolds K, et al. Global Disparities of Hypertension Prevalence and Control: A Systematic Analysis of Population-Based Studies From 90 Countries. Circulation. 2016;134(6):441–50 DOI: 10.1161/CIRCULATIONAHA.115.018912.

5. Mills KT, Stefanescu A, He J. The global epidemiology of hypertension. Nat Rev Nephrol. 2020;16(4):223–37 DOI: 10.1038/s41581-019-0244-2.

6. Akinlua JT, Meakin R, Umar AM, Freemantle N. Current Prevalence Pattern of Hypertension in Nigeria: A Systematic Review. PLoS One. 2015;10(10):e0140021 DOI: 10.1371/journal.pone.0140021.

7. Murthy GV, Fox S, Sivasubramaniam S, Gilbert CE, Mahdi AM, Imam AU, et al. Prevalence and risk factors for hypertension and association with ethnicity in Nigeria: results from a national survey. Cardiovasc J Afr. 2013;24(9-10):344–50 DOI: 10.5830/CVJA-2013-058.

8. Mozaffarian D, Fahimi S, Singh GM, Micha R, Khatibzadeh S, Engell RE, et al. Global sodium consumption and death from cardiovascular causes. N Engl J Med. 2014;371(7):624–34 DOI: 10.1056/NEJMoa1304127.

9. Oyebode O, Oti S, Chen YF, Lilford RJ. Salt intakes in sub-Saharan Africa: a systematic review and meta-regression. Popul Health Metr. 2016;14:1 DOI: 10.1186/s12963-015-0068-7.

10. Collaborators GBDD. Health effects of dietary risks in 195 countries, 1990-2017: a systematic analysis for the Global Burden of Disease Study 2017. Lancet. 2019;393(10184):1958–72 DOI: 10.1016/S0140-6736(19)30041-8.

11. Schwingshackl L, Schwedhelm C, Hoffmann G, Knuppel S, Iqbal K, Andriolo V, et al. Food Groups and Risk of Hypertension: A Systematic Review and Dose-Response Meta-Analysis of Prospective Studies. Adv Nutr. 2017;8(6):793–803 DOI: 10.3945/an.117.017178.

12. Batubo NP, Moore JB, Zulyniak MA. Dietary factors and hypertension risk in West Africa: a systematic review and meta-analysis of observational studies. J Hypertens. 2023;41(9):1376–88 DOI: 10.1097/HJH.0000000000003499.

13. Ojji D, Nigeria Sodium Study T. Developing long-term strategies to reduce excess salt consumption in Nigeria. Eur Heart J. 2022;43(13):1277–9 DOI: 10.1093/eurheartj/ehac025.

14. Lee RD, Nieman DC. Nutritional assessment. 6 ed: McGraw-Hill, New York, NY,; 2013.

15. Meybeck A, Redfern S, Paoletti F, Strassner C, Office of Assistant Director-General (Agriculture Department). Assessing sustainable diets within the sustainability assessment of food and agriculture systems. Mediterranean diet, organic food: new challenges: FAO; 2015.

16. Goulet J, Nadeau G, Lapointe A, Lamarche B, Lemieux S. Validity and reproducibility of an interviewer-administered food frequency questionnaire for healthy French-Canadian men and women. Nutr J. 2004;3:13 DOI: 10.1186/1475-2891-3-13.

17. Ene-Obong HN, Onuoha NO, Aburime LU, Emebu PK. Prevalence of overweight/obesity and hypertension amongst adults in three ethnic groups in Nigeria. Asia Journal of Public Health. 2013;2:12–9.

18. Chan AW, Tetzlaff JM, Altman DG, Laupacis A, Gotzsche PC, Krleza-Jeric K, et al. SPIRIT 2013 statement: defining standard protocol items for clinical trials. Ann Intern Med. 2013;158(3):200–7 DOI: 10.7326/0003-4819-158-3-201302050-00583.

19. Adebamowo SN, Eseyin O, Yilme S, Adeyemi D, Willett WC, Hu FB, et al. A Mixed-Methods Study on Acceptability, Tolerability, and Substitution of Brown Rice for White Rice to Lower Blood Glucose Levels among Nigerian Adults. Front Nutr. 2017;4:33 DOI: 10.3389/fnut.2017.00033.

20. Mertens E, Kuijsten A, Geleijnse JM, Boshuizen HC, Feskens EJM, Van’t Veer P. FFQ versus repeated 24-h recalls for estimating diet-related environmental impact. Nutr J. 2019;18(1):2 DOI: 10.1186/s12937-018-0425-z.

21. Steinemann N, Grize L, Ziesemer K, Kauf P, Probst-Hensch N, Brombach C. Relative validation of a food frequency questionnaire to estimate food intake in an adult population. Food Nutr Res. 2017;61(1):1305193 DOI: 10.1080/16546628.2017.1305193.

22. Willett W. Foreword. The validity of dietary assessment methods for use in epidemiologic studies. Br J Nutr. 2009;102 Suppl 1:S1–2 DOI: 10.1017/S0007114509993102.

23. Swinscow T.D.V., Campbell M.J. Statistics at square one. Correlation and regression BMJ [19:[Available from: https://www.bmj.com/about-bmj/resources-readers/publications/statistics-square-one/11-correlation-and-regression, https://www.bmj.com/about-bmj/resources-readers/publications/statistics-square-one/11-correlation-and-regression.

24. Faul F, Erdfelder E, Buchner A, Lang AG. Statistical power analyses using G*Power 3.1: tests for correlation and regression analyses. Behav Res Methods. 2009;41(4):1149–60 DOI: 10.3758/BRM.41.4.1149.

25. Gupta KK, Attri JP, Singh A, Kaur H, Kaur G. Basic concepts for sample size calculation: Critical step for any clinical trials! Saudi J Anaesth. 2016;10(3):328–31 DOI: 10.4103/1658-354X.174918.

26. Thabane L, Ma J, Chu R, Cheng J, Ismaila A, Rios LP, et al. A tutorial on pilot studies: the what, why and how. BMC Med Res Methodol. 2010;10:1 DOI: 10.1186/1471-2288-10-1.

27. Federal Ministry of Health. National Nutritional Guideline On Non-Communicable Disease Prevention, Control and Management In: Federal Ministry of Health A, editor. 2014. p. 62, https://extranet.who.int/countryplanningcycles/sites/default/files/planning_cycle_repository/nigeria/nutritionalguideline.pdf.

28. Ministry of Health RoG. Strategy for the Management, Prevention and Control of Chronic Non-Communicable Diseases in Ghana. August, 2012, chrome-extension://efaidnbmnnnibpcajpcglclefindmkaj/https://extranet.who.int/nutrition/gina/sites/default/filesstore/GHA-2012-NCDs.pdf.

29. U.S. Department of Agriculture. AMPM - USDA Automated Multiple-Pass Method 2021 [Available from: https://www.ars.usda.gov/northeast-area/beltsville-md-bhnrc/beltsville-human-nutrition-research-center/food-surveys-research-group/docs/ampm-usda-automated-multiple-pass-method/, https://www.ars.usda.gov/northeast-area/beltsville-md-bhnrc/beltsville-human-nutrition-research-center/food-surveys-research-group/docs/ampm-usda-automated-multiple-pass-method/.

30. Blanton CA, Moshfegh AJ, Baer DJ, Kretsch MJ. The USDA Automated Multiple-Pass Method accurately estimates group total energy and nutrient intake. J Nutr. 2006;136(10):2594–9 DOI: 10.1093/jn/136.10.2594.

31. Conway JM, Ingwersen LA, Moshfegh AJ. Accuracy of dietary recall using the USDA five-step multiple-pass method in men: an observational validation study. J Am Diet Assoc. 2004;104(4):595–603 DOI: 10.1016/j.jada.2004.01.007.

32. Fatihah F, Ng BK, Hazwanie H, Norimah AK, Shanita SN, Ruzita AT, et al. Development and validation of a food frequency questionnaire for dietary intake assessment among multi-ethnic primary school-aged children. Singapore Med J. 2015;56(12):687–94 DOI: 10.11622/smedj.2015190.

33. Shapiro SS, Wilk MB. An analysis of variance test for normality (complete samples)†. Biometrika. 1965;52(3-4):591–611 DOI: 10.1093/biomet/52.3-4.591.

34. Massey FJ. The Kolmogorov-Smirnov Test for Goodness of Fit. Journal of the American Statistical Association. 1951;46(253):68–78 DOI: 10.1080/01621459.1951.10500769.

35. R Core Team. A language and environment for statistical computing. R Foundation for Statistical Computing. 4.3.1 ed: Vienna, Austria; 2020, https://www.R-project.org/.

36. Cade J, Thompson R, Burley V, Warm D. Development, validation and utilisation of food-frequency questionnaires - a review. Public Health Nutr. 2002;5(4):567–87 DOI: 10.1079/PHN2001318.

37. Rohrmann S, Klein G. Validation of a short questionnaire to qualitatively assess the intake of total fat, saturated, monounsaturated, polyunsaturated fatty acids, and cholesterol. J Hum Nutr Diet. 2003;16(2):111–7 DOI: 10.1046/j.1365-277x.2003.00425.x.

38. Rey D, Neuhäuser M. Wilcoxon-Signed-Rank Test. In: Lovric M, editor. International Encyclopedia of Statistical Science;10.1007/978-3-642-04898-2_616 10.1007/978-3-642-04898-2_616. Berlin, Heidelberg: Springer Berlin Heidelberg; 2011. p. 1658–9 DOI: 10.1007/978-3-642-04898-2_616.

39. Bland JM, Altman DG. Statistical methods for assessing agreement between two methods of clinical measurement. Lancet. 1986;1(8476):307–10, https://www.ncbi.nlm.nih.gov/pubmed/2868172.

40. Savija N, Leong DP, Pinthus J, Karampatos S, Shayegan B, Mian R, et al. Development and Comparability of a Short Food-Frequency Questionnaire to Assess Diet in Prostate Cancer Patients: The Role of Androgen Deprivation Therapy in CArdiovascular Disease - A Longitudinal Prostate Cancer Study (RADICAL PC) Substudy. Curr Dev Nutr. 2021;5(11):nzab106 DOI: 10.1093/cdn/nzab106.

41. Koo TK, Li MY. A Guideline of Selecting and Reporting Intraclass Correlation Coefficients for Reliability Research. J Chiropr Med. 2016;15(2):155–63 DOI: 10.1016/j.jcm.2016.02.012.

42. Challa HJ AM, Uppaluri KR,. DASH Diet To Stop Hypertension: In: StatPearls [Internet]. Treasure Island (FL): StatPearls Publishing; January 23, 2023 [Available from: https://www.ncbi.nlm.nih.gov/books/NBK482514/, https://www.ncbi.nlm.nih.gov/books/NBK482514/.

43. Streppel MT, de Vries JH, Meijboom S, Beekman M, de Craen AJ, Slagboom PE, et al. Relative validity of the food frequency questionnaire used to assess dietary intake in the Leiden Longevity Study. Nutr J. 2013;12:75 DOI: 10.1186/1475-2891-12-75.

44. Marks GC, Hughes MC, van der Pols JC. Relative validity of food intake estimates using a food frequency questionnaire is associated with sex, age, and other personal characteristics. J Nutr. 2006;136(2):459–65 DOI: 10.1093/jn/136.2.459.

45. Wong JE, Parnell WR, Black KE, Skidmore PM. Reliability and relative validity of a food frequency questionnaire to assess food group intakes in New Zealand adolescents. Nutr J. 2012;11:65 DOI: 10.1186/1475-2891-11-65.

46. Yun SH, Choi B-Y, Kim M-K. The Effect of Seasoning on the Distribution of Nutrient Intakes by a Food-Frequency Questionnaire in a Rural Area. Korean J Nutr. 2009;42(3):246–55, 10.4163/kjn.2009.42.3.246.

47. Aoun C, Bou Daher R, El Osta N, Papazian T, Khabbaz LR. Reproducibility and relative validity of a food frequency questionnaire to assess dietary intake of adults living in a Mediterranean country. PLoS One. 2019;14(6):e0218541 DOI: 10.1371/journal.pone.0218541.

48. Willett WC, Sampson L, Stampfer MJ, Rosner B, Bain C, Witschi J, et al. Reproducibility and validity of a semiquantitative food frequency questionnaire. Am J Epidemiol. 1985;122(1):51–65 DOI: 10.1093/oxfordjournals.aje.a114086.

49. Fallaize R, Forster H, Macready AL, Walsh MC, Mathers JC, Brennan L, et al. Online dietary intake estimation: reproducibility and validity of the Food4Me food frequency questionnaire against a 4-day weighed food record. J Med Internet Res. 2014;16(8):e190 DOI: 10.2196/jmir.3355.

50. Baranowski T, Willett W. 4924-Hour Recall and Diet Record Methods. Nutritional Epidemiology;10.1093/acprof:oso/9780199754038.003.0004 10.1093/acprof:oso/9780199754038.003.0004: Oxford University Press; 2012. p. 0 DOI: 10.1093/acprof:oso/9780199754038.003.0004.

51. Lee RD, Nieman DC, Rainwater M. Comparison of eight microcomputer dietary analysis programs with the USDA Nutrient Data Base for Standard Reference. J Am Diet Assoc. 1995;95(8):858–67 DOI: 10.1016/S0002-8223(95)00240-5.

52. Batubo NP, Nwanze NM, Alikor CA, Auma CI, Moore JB, Zulyniak MA. Empowering Healthcare Professionals in West Africa. A Feasibility Study and Qualitative Assessment of a Dietary Screening Tool to Identify Adults at High Risk of Hypertension. 2023; 10.1101/2023.11.09.23297914 https://doi.org/10.1101/2023.11.09.23297914 DOI: 10.1101/2023.11.09.23297914.

53. Gibson RS, Charrondiere UR, Bell W. Measurement Errors in Dietary Assessment Using Self-Reported 24-Hour Recalls in Low-Income Countries and Strategies for Their Prevention. Adv Nutr. 2017;8(6):980–91 DOI: 10.3945/an.117.016980.

